# Evolving landscape of economic evaluations of HIV pre-exposure prophylaxis and pre-exposure prophylaxis implementation strategies: A systematic review

**DOI:** 10.1101/2024.12.02.24318351

**Authors:** Min Xi, Darrell H. S. Tan, Stefan D. Baral, Howsikan Kugathasan, Lisa Masucci, Becky Skidmore, Derek R. MacFadden, Kednapa Thavorn, Sharmistha Mishra

## Abstract

**Introduction:** Economic evaluations of HIV pre-exposure prophylaxis (PrEP) and associated implementation strategies guide evidence-based policies, programmes, and resource allocation. Since 2015, there has been an evolution in PrEP modalities, implementation strategies, and prioritization of key populations with unmet HIV prevention needs, alongside the scale-up of other HIV prevention interventions. Our systematic review describes the evolving landscape of economic evaluations of PrEP to help identify evidence gaps relevant to the current HIV epidemic and response (PROSPERO: <u>CRD42016038440</u>).

**Methods:** We searched five databases, without language restrictions, for peer-reviewed economic evaluations from inception to December 21, 2024. We describe the evolution of study characteristics over time, including perspective of analysis, region, population, PrEP modality/implementation strategy, and comparators.

**Results:** Of 5,046 studies identified, 114 met inclusion criteria, of which 81 examined HIV epidemics in 2015 or later and 16 adopted a societal perspective. HIV epidemics studied primarily spanned countries in Sub-Saharan Africa (N=45) and in North America (N=32). Modeled populations for receipt of PrEP primarily comprised: gay, bisexual, and other men who have sex with men (N=66), female sex workers (N=23), serodifferent partnerships (N=16), and persons who inject drugs (N=12). Most evaluated oral, daily PrEP (N=70), followed by on-demand PrEP (N=12), long-acting injectable PrEP (N=12), and others (e.g., vaginal ring, topical gel; N=7). Ten studies compared different PrEP modalities with each other. Three studies evaluated different implementation strategies to increase PrEP uptake, adherence, and persistence. Of the 108 studies that compared PrEP to a combination of other HIV prevention interventions, only 25 scaled up at least part of the comparator over time.

**Discussion:** To support decision-making, future economic evaluations should consider costs and benefits beyond the health system and consider comparators that better reflect the current HIV response across regions and populations. The increasing availability of novel PrEP modalities allows future studies to evaluate a mix of PrEP modalities and person-centered implementation strategies.

**Conclusions:** The growing number of PrEP economic evaluations have not kept pace with emerging PrEP modalities or the current HIV epidemic/response, resulting in challenges in making evidence-based policies, programmes, and resource allocation.

## Introduction

The last decade has seen an expansion in the adoption and roll-out of oral pre-exposure prophylaxis (PrEP), consisting of emtricitabine and either tenofovir disoproxil fumarate or tenofovir alafenamide, for HIV prevention. Oral PrEP was first authorized for use in the United States in 2012 [1]. There has also been an expansion in PrEP modalities (daily or “on-demand” oral [2]; long-acting injectable and topical formulations [3]), and implementation strategies (e.g., mobile phone applications [4] or online modules [5] with information on PrEP or peer support for PrEP initiation and persistence). By 2015, PrEP became a key component of combination HIV prevention globally when the World Health Organization recommended oral PrEP for individuals at “substantial risk” of acquiring HIV (annual HIV risk greater than 3%) [6]. By 2021, 144 countries recommended oral PrEP within their national HIV prevention guidelines, with 14 more planning to adopt PrEP into their guidelines within the next two years [7].

PrEP implementation strategies have been a central focus for HIV programmes in the last decade [8, 9] because, despite the inclusion of PrEP in national guidelines, coverage remains low in many countries [10]. PrEP use in 2020 was at 8% of the 2025 global target set by UNAIDS [11]. While individual-level efficacy in preventing HIV acquisition is high (upwards of 99% [12]), real-world individual-level or partnership-level effectiveness has been lower [13–16] partly due to challenges with adherence and persistence [17, 18]. Although mathematical modelling studies suggest large potential for reducing sexual transmission of HIV at the population-level, such outcomes depend on PrEP reaching individuals with the most unmet prevention needs who are part of sexual networks with high rates of HIV transmission [19–21].

Economic evaluations of PrEP and PrEP implementation strategies are valuable for informing policy decisions, optimizing resource allocation, and guiding the adoption of different implementation strategies [22]. To date, there have been 12 reviews of economic evaluations of PrEP [23–34], but they were largely restricted with respect to geography and/or populations. Two reviews synthesized evidence from a single country (United States) [23, 24], three were restricted to a subset of countries (high-income, early PrEP adopting countries) [32, 34] or to one region (Sub-Saharan Africa) [33], and two focused on specific populations (gay, bisexual, and other men who have sex with men; persons who inject drugs) [23, 24]. These earlier reviews were also restricted to economic evaluations conducted prior to the recent expansions in PrEP roll-out and expansions in PrEP modalities and implementation strategies [23–30].

The evolution of PrEP roll-out over the last decade signals a need to characterize how economic evaluations may have evolved (e.g., the examination of different PrEP modalities and implementation strategies) to keep pace with implementation-relevant questions for decision-makers and communities most affected by HIV [9]. The most recent systematic review, which included economic evaluations conducted between 2010 and 2020, was not designed to describe the evolution of comparators used in PrEP economic evaluations over time and whether these comparators reflected the evolving HIV epidemic response [31] (e.g., implementation and scale-up of concurrent HIV prevention tools, strategies, and guideline [35–37]). For example, comparison of PrEP to steady levels of other modes of primary and secondary HIV prevention at a population level may be less applicable for decision-making in recent years, especially given acceleration in sustained viral suppression as secondary HIV prevention [38]. The scale-up of combination HIV prevention interventions has led to large reductions in HIV incidence globally, including in countries with some of the highest burden of HIV, and incidence reductions in populations at highest risk of HIV acquisition (including gay, bisexual, and other men who have sex with men, cisgender female sex workers and other women engaged in sex work, transgender women, persons who inject drugs) [39]. Therefore, the questions being asked by decision-makers involve comparing and choosing between different PrEP modalities and implementation strategies [40]. In doing so, decision-makers seek strategies to improve each step of the PrEP cascade [40] (including persistence [18]) and determine where and among whom new HIV infections continue to be acquired despite local efforts in scaling up combination HIV prevention [39].

In this study, we sought to: 1) describe the evolving landscape of economic evaluations of PrEP and its implementation strategies over time and across epidemic contexts (population focus, country/region) and 2) highlight gaps in the literature. Our goal was to provide guidance for future economic evaluations (e.g., which PrEP interventions/implementation strategies to evaluate and which comparators, HIV epidemic context, and perspective should be used during analyses) to better support decision-making in the evolving landscape of PrEP implementation.

## Methods

Our systematic review followed the Preferred Reporting items for Systematic Reviews and Meta-Analyses (PRISMA) guidelines (Appendix A) [41]. Our full review protocol was peer-reviewed and published [42] and registered in PROSPERO (registration number <u>CRD42016038440</u>). Our full review protocol objectives included: 1) estimating the incremental cost per health outcome of PrEP or PrEP implementation strategies compared to placebo, status quo, or other HIV prevention strategies, 2) assessing the variability in and quality of economic evaluations of PrEP or PrEP implementation strategies, and 3) identifying potential sources of heterogeneity in the cost-effectiveness of PrEP or PrEP implementation strategies. In this first paper, we described the trends and characteristics of PrEP economic evaluations over time and across epidemic contexts, with a focus on relevance to evolving decision-making needs in the current stage of the global HIV epidemic - specifically, regarding PrEP modalities, implementation strategies, comparators [39].

### Search Strategy and Inclusion/Exclusion Criteria

An information specialist (BS) developed the strategy in consultation with the review team. The strategies were executed in Ovid MEDLINE® ALL, Embase Classic+Embase (Ovid), and the NHS Economic Evaluation Database (Wiley). An initial search was conducted on August 30, 2019, updated to December 23, 2021, and updated again to December 21, 2024. The NHS Economic Evaluation Database was not included in the updated search as it was no longer being updated and was removed from the Cochrane Library. The search included a combination of controlled vocabulary (e.g., “HIV Infections/pc [Prevention & Control]”, “Pre-Exposure Prophylaxis”, “Models, Economic”) and free-text terms (e.g., “HIV prevention”, “PrEP”, “economic evaluation”). There were no language or date restrictions but, where possible, animal-only records and conference abstracts were removed from the results. The full search strategy is available in Appendix B. We (MX) also conducted a bibliographic hand search of relevant reviews to identify studies that may have been missed by the electronic database searches.

We included full economic evaluations (i.e., cost-minimization analyses, cost-benefit analyses, cost-effectiveness analyses, or cost-utility analyses) that compared both costs and outcomes of PrEP or of a PrEP implementation strategy [43]. Studies could compare PrEP as an intervention to placebo, to other HIV prevention interventions (e.g., ART, condoms, HIV testing, HIV counselling, male circumcision), or to other PrEP modalities; and/or compare different PrEP implementation strategies to each other. We excluded editorials, conference abstracts, and review articles. We excluded studies evaluating emtricitabine and either tenofovir disoproxil fumarate or tenofovir alafenamide for post-exposure prophylaxis rather than PrEP. We excluded economic evaluations focussed solely on the prevention of parent-to-child transmission of HIV.

Following duplicate removal, titles and abstracts were screened independently by two reviewers (two of either MX, KT, SM) for inclusion/exclusion using Covidence (Veritas Health Innovation, Melbourne, Australia). For included titles/abstracts and in cases where the title and abstract were deemed insufficient for an exclusion decision, two reviewers (two of either MX, HK, KT, SM) independently conducted full text screening against the inclusion/exclusion criteria. Discrepancies during title and abstract and full text screening were resolved by consensus.

### Data extraction

Data from the included publications were extracted by one reviewer (MX, LM, HK, or NM) and verified by a second reviewer (MX, LM, HK, NM, or OD). For study purposes, we categorized the extracted variables as follows: (1) epidemic context; (2) perspective of analyses; (3) intervention, and (4) comparator (Appendix C).

Epidemic context pertained to the underlying epidemic and population under study, with variables including country and population who received PrEP and populations included in the outcome measures. Variables related to the underlying epidemic included: country and World Bank region[44], publication year and model time-horizon, HIV epidemic period modelled, and calendar time-period of PrEP use. Populations included population focus, such as gay, bisexual, and other men who have sex with men, persons who inject drugs, female sex workers, male sex workers, HIV-serodifferent partners, adolescent girls and young women; and the wider population (e.g., total adult population).

For the perspective of analyses, we extracted data on whether evaluations were conducted using a health system, societal perspective, and/or other perspective. Intervention variables included: concurrent interventions (e.g., concomitant ART, condoms, etc.); PrEP modality (oral [daily or on-demand], continuous, long-acting, vaginal ring, topical gel]); PrEP implementation strategies (e.g., PrEP provider strategy, strategies to increase PrEP coverage and/or to improve adherence to PrEP).

For comparator-related variables, we extracted the comparator type (e.g., ART alone; ART with other HIV prevention interventions excluding PrEP; other prevention interventions without ART). If PrEP modality or implementation strategies were evaluated, then comparators could also include: alternate PrEP modalities with or without ART and/or other prevention interventions; alternate PrEP implementation strategies with or without ART and/or other prevention interventions. If comparators did not fall into the above categories, we classified them as “other” and provided details in the extraction form. Finally, we extracted data on whether any of the interventions included in the comparator was scaled up during the period of comparative analyses (e.g., increased coverage ART).

### Data Synthesis

We conducted a narrative synthesis of economic evaluations and summarized the results in tabular and graphical formats (performed in R version 4.2.0 [45]). We described the following characteristics of the included studies: epidemic context, perspective of analysis, PrEP modality or implementation strategy, and comparator(s). We examined whether comparators used in analyses included the use and scale up of various modes of primary and secondary population-level HIV prevention interventions [35, 36, 38]. Specifically, we quantified the number of studies that included ART or other HIV prevention interventions in the comparator and whether models accounted for the increased uptake/coverage of at least one of these HIV prevention interventions over time. For studies that compared one PrEP modality or implementation strategy to another, we described the PrEP modality, HIV prevention interventions, and/or PrEP implementation strategies included in the comparator. Results were stratified by year of publication to show the evolution of the landscape of PrEP and PrEP implementation economic evaluation studies over time. We also quantified the number of studies published in 2015 or later, following World Health Organization endorsement of PrEP [6].

## Results

### Study selection

Figure 1 describes the study inclusion process. Our search identified 5,046 unique records. Following title and abstract screening, 295 studies were retrieved for full text screening. 114 studies met the inclusion criteria and were included in our review.

**Figure 1.**
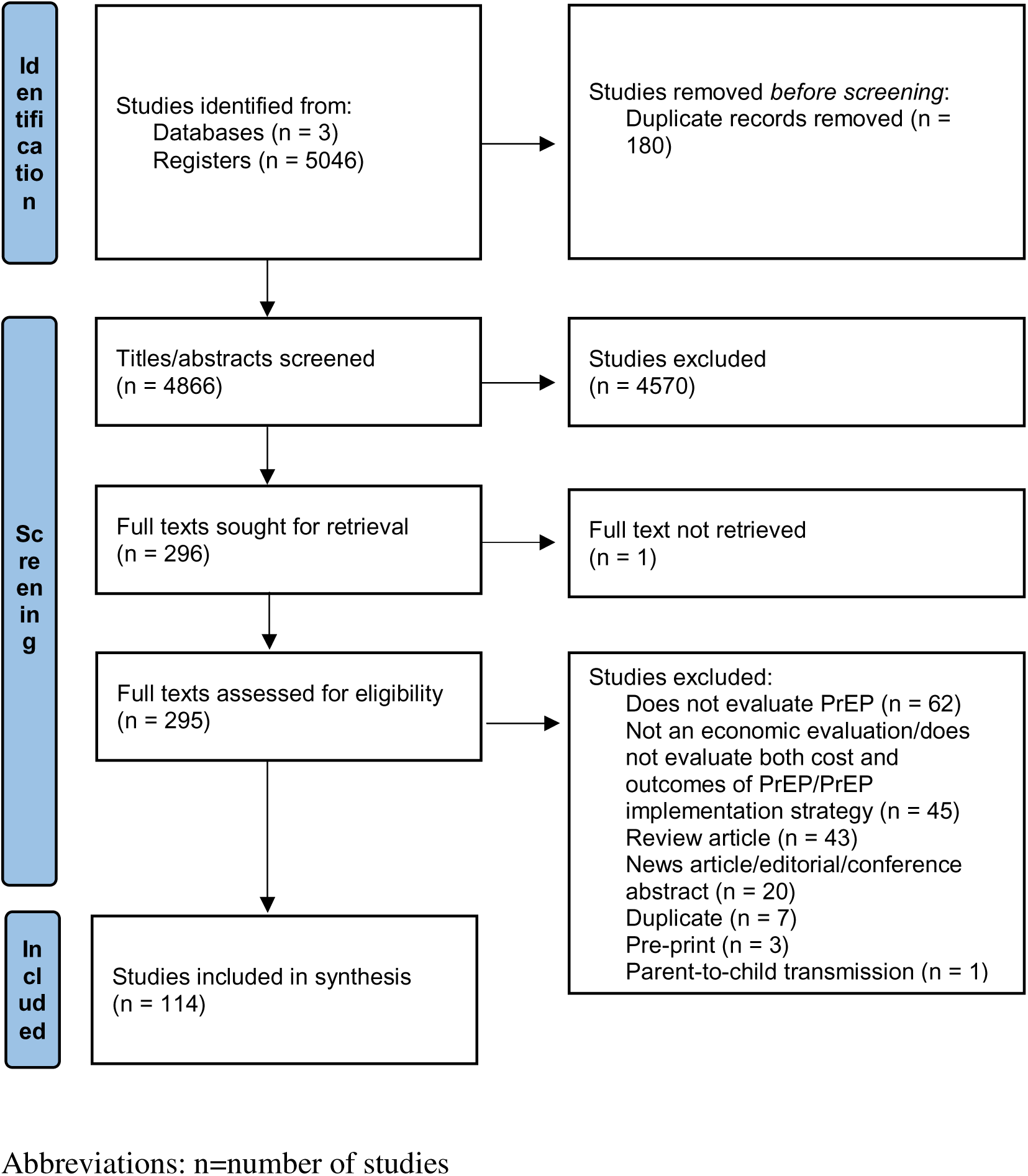
PRISMA flow diagram of study selection.

### Timeline in number of economic evaluations by key events

We identified an increasing trend in the number of economic evaluations that assessed the cost-effectiveness of PrEP in recent years. Six studies (5%) [46–51] were published between 2008 and 2011 prior to US FDA approval of PrEP [1], 15 studies (13%) [52–66] were published between 2012 and 2014 following US FDA approval of PrEP [1], 20 studies (18%) [67–86] were published between 2015 and 2016 following WHO endorsement of PrEP [6], and 73 studies (64%) [4, 87–158] were published between 2017 and 2024 following the availability of generic PrEP [159, 160].

Eighty-one studies (71%) [4, 49, 50, 53–63, 67, 68, 71–73, 75, 78–81, 84, 85, 88–92, 94, 98, 102–113, 117–124, 128–144, 146, 147, 149–155, 157, 158] simulated HIV epidemics that included a time-period in 2015 or later, with 59 studies [50, 53, 60, 62, 63, 71, 72, 75, 78–81, 84, 85, 88–90, 92, 94, 98, 102, 106, 109–113, 118–123, 128, 129, 131–133, 135–144, 146, 147, 149–155, 157, 158] simulating epidemics and outcomes in 2030 or later (up to 2103 [136]). One study simulated an HIV epidemic prior to 2015 [47]. The remaining 32 (28%) studies did not report the year(s) of HIV epidemic captured in the model [46, 48, 51, 52, 64–66, 69, 70, 74, 76, 77, 82, 83, 86, 87, 93, 95–97, 99–101, 114–116, 125–127, 145, 148, 156].

### Distribution of PrEP economic evaluations by perspective of analysis and year of publication

Ninety-four of the 114 (82%) included studies conducted analyses from a health system perspective [4, 47, 49–51, 54–63, 66–69, 71, 73–83, 86–88, 90–92, 94–100, 102–107, 109–112, 114, 116–126, 128–130, 132–141, 143–150, 152–158]. These studies captured healthcare costs and associated health outcomes within the context of the health system (e.g., costs covered by a public healthcare payer). From 2013 to 2024 (i.e., following the first authorized use of PrEP by the US FDA), 71% to 100% of PrEP cost-effectiveness studies each year were conducted from a health system perspective (Figure 2).

**Figure 2.**
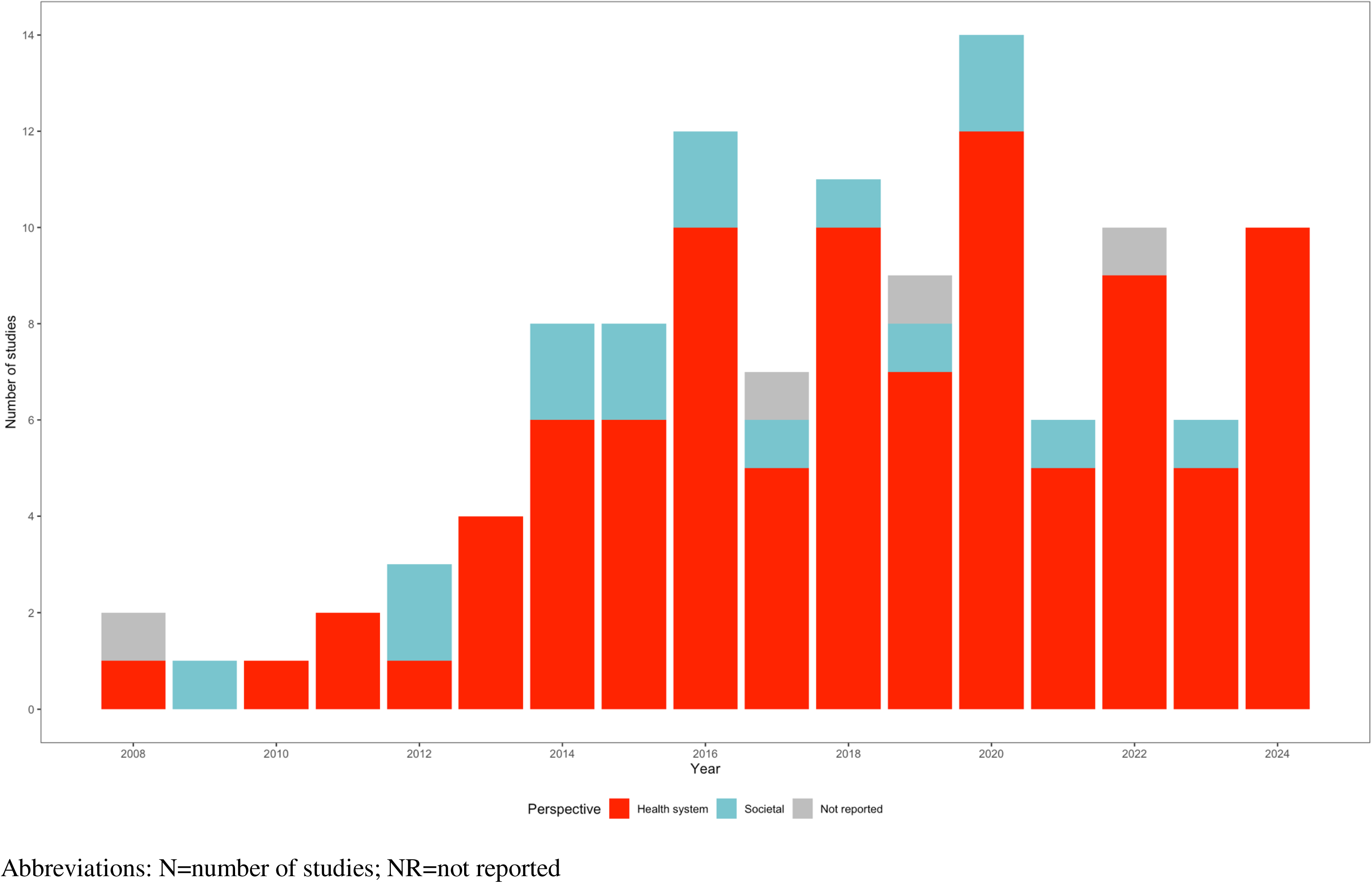
Number of PrEP cost-effectiveness studies by perspective of the analysis and year of publication (N=114)

Less than a fifth of included studies conducted analyses from a societal perspective (n=16, n=14%) and examined the full range of costs and outcomes associated with PrEP use including productivity costs [48, 52, 53, 64, 65, 70, 72, 84, 85, 89, 101, 108, 115, 127, 131, 142]. Four studies did not report the perspective taken for analysis [46, 93, 113, 151].

### Economic evaluation of PrEP by region, population who received PrEP, and populations included in outcome measures

Forty-five (39%) studies modeled HIV epidemics in countries across in Sub-Saharan Africa (Figure 3a) [49–52, 55–60, 62, 66–69, 71, 73–75, 77, 79–81, 84, 88, 90, 91, 100, 106, 110, 111, 117, 118, 123, 125, 129, 133, 137, 139, 141, 143, 146, 149, 150, 155]. Almost a third of the eighty-seven included studies modeled HIV epidemics in North America (n=32, 28%) [4, 47, 48, 53, 63, 64, 72, 76, 82, 83, 85–87, 89, 93, 96, 97, 102, 107, 116, 120–122, 126, 127, 132, 134, 135, 138, 145, 147, 153]. The remaining studies modeled HIV epidemics in Europe and Central Asia (n=14, 12%) [60, 65, 70, 78, 92, 94, 95, 101, 112, 124, 128, 130, 136, 142], East Asia and Pacific (n=17, 15%) [60, 61, 98, 103, 104, 108, 113, 114, 131, 140, 144, 151, 152, 154, 156–158], Latin America and Caribbean (n=5, 4%) [54, 60, 99, 105, 148], South Asia (n=4, 3%) [60, 109, 119, 151], and Middle East and North Africa (n=1, 1%) [115].

**Figure 3a.**
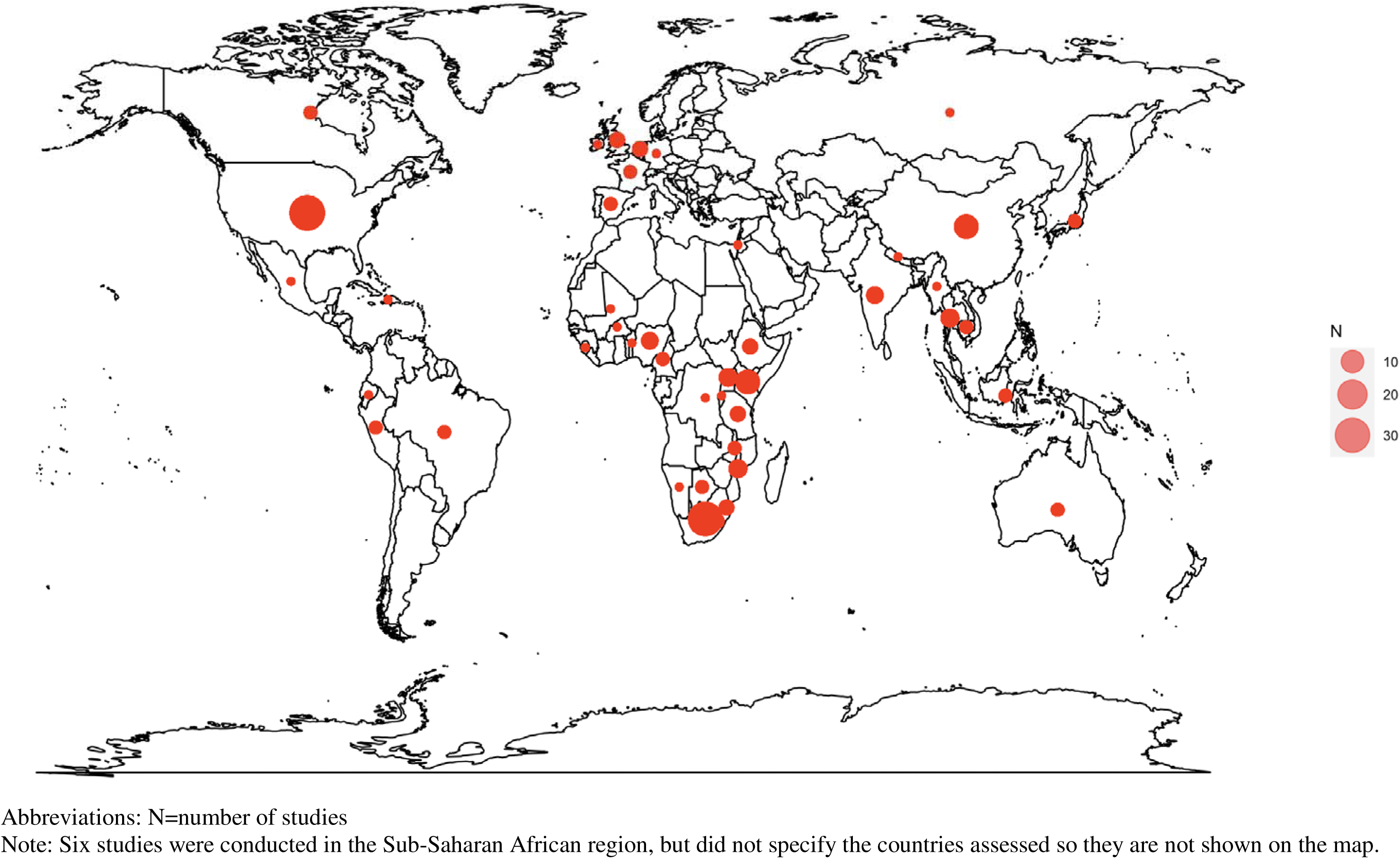
Geographic location of PrEP intervention and implementation strategy cost-effectiveness models (N=114)

**Figure 3b.**
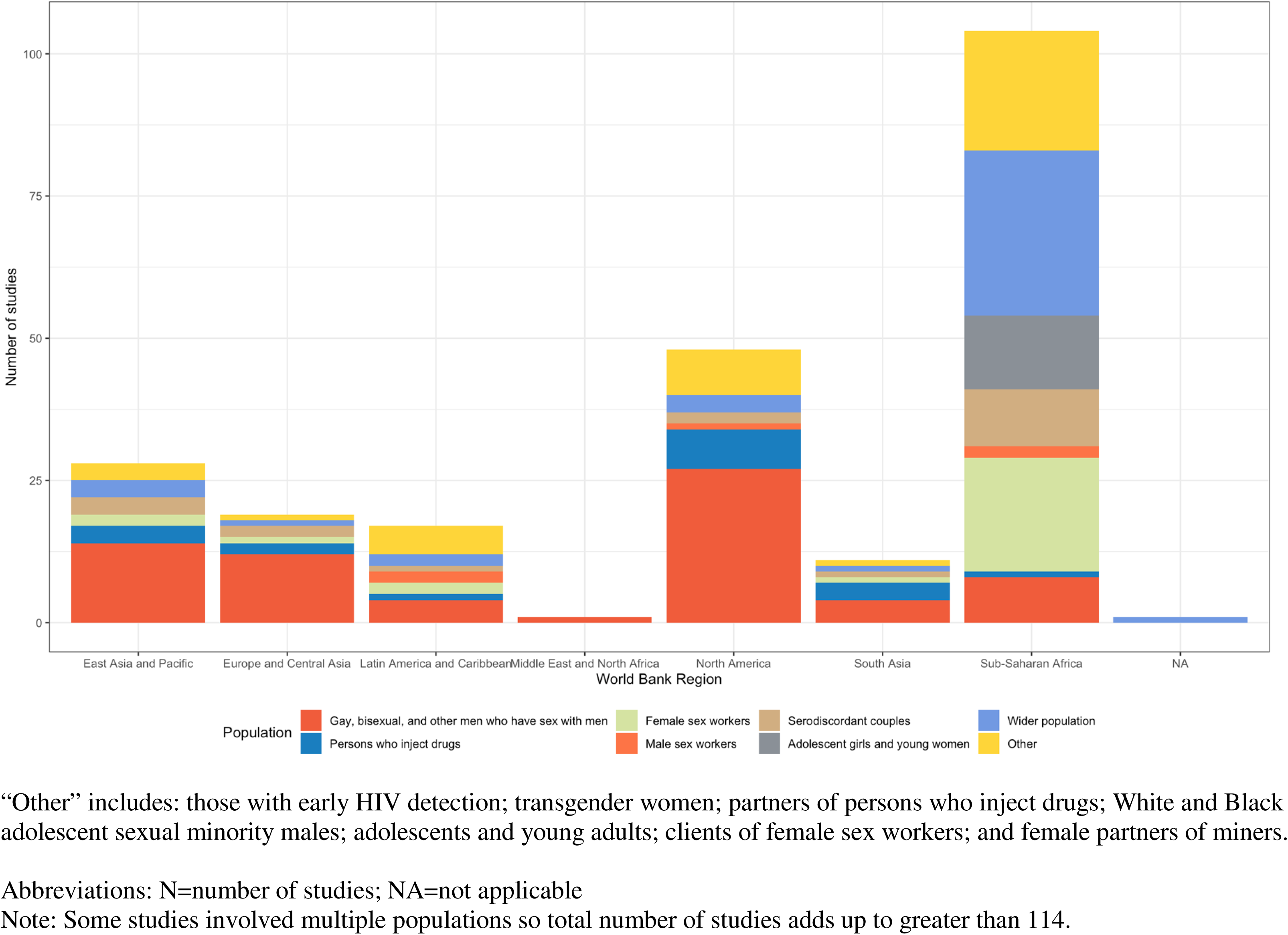
Number of studies by World Bank region and study population who received PrEP (N=114)

Except in the context of HIV epidemics across Sub-Saharan Africa, most studies modelled the receipt and use of PrEP among gay, bisexual, and other men who have sex with men across HIV epidemics across each of the World Bank regions: 27 (84%) in North America [4, 47, 48, 53, 63, 64, 72, 76, 82, 83, 85–87, 102, 107, 116, 120–122, 126, 132, 134, 135, 138, 145, 147, 153], 12 (86%) in Europe and Central Asia [60, 78, 92, 94, 95, 101, 112, 124, 128, 130, 136, 142], 14 (82%) in east Asia and Pacific [60, 61, 98, 103, 104, 108, 113, 114, 140, 144, 151, 156–158], four (80%) in Latin America and Caribbean [54, 60, 99, 148], four (100%) in South Asia [60, 109, 119], and one (100%) in Middle East and North Africa [115] (Figure 3b).

In contrast, nearly two-thirds (n=31, 69%) of the studies of HIV epidemics Sub-Saharan Africa countries examined PrEP use in the wider population, defined as the total adult male and/or female population in a region (Figure 3b) [49, 50, 52, 56, 58–60, 62, 66, 67, 74, 75, 77, 80, 81, 88, 90, 100, 106, 110, 111, 117, 118, 129, 137, 139, 141, 143, 146, 149, 155]. In studies modelling HIV epidemics across Sub-Saharan Africa, PrEP receipt and use was also assessed in the following populations: female sex workers (n=21, 47%) [60, 75, 80, 81, 84, 90, 91, 100, 111, 117, 118, 123, 129, 133, 139, 141, 143, 146, 149, 150, 155], male sex workers (n=2, 4%) [91, 146], HIV-serodifferent partners (n=11, 22%) [51, 55, 57, 60, 68, 69, 71, 73, 84, 123, 143], adolescent girls and young women (n=13, 29%) [52, 74, 88, 90, 100, 111, 117, 123, 129, 139, 143, 146, 155], gay, bisexual, and other men who have sex with men (n=9, 20%) [59, 60, 80, 81, 91, 118, 139, 141, 150], and persons who inject drugs (n=2, 4%) [60, 141].

Across the 114 included studies, particularly studies modelling World Bank regions other than Sub-Saharan Africa, few studies assessed the cost-effectiveness of PrEP among other key populations including HIV-serodifferent partners (n=14, 15%) [51, 55, 57, 60, 63, 68–71, 73, 84, 97, 123, 131, 143, 154], persons who inject drugs (n=12, 11%) [60, 63, 65, 89, 93, 96, 109, 119–121, 141, 147], female sex workers (n=23, 51%) [60, 75, 80, 81, 84, 90, 91, 100, 111, 117, 118, 123, 129, 133, 139, 141, 143, 146, 148–150, 155, 157], male sex workers (n=5, 4%) [54, 91, 116, 146, 148], and adolescent girls and young women (n=13, 11%) [52, 74, 88, 90, 100, 111, 117, 123, 129, 139, 143, 146, 155].

Overall, 94 studies (82%) [4, 46–63, 65–71, 73–86, 88–91, 94, 96–99, 102–124, 127, 129–133, 135, 136, 138–143, 145–147, 149, 150, 152, 153, 155] included transmission dynamics (i.e., indirect benefit to partners of people who received PrEP) and 20 (18%) [64, 72, 87, 92, 93, 95, 100, 101, 125, 126, 128, 134, 137, 144, 148, 151, 154, 156–158] did not.

### Economic evaluations by PrEP modality

Most studies (n= 91, 80%) assessed oral PrEP, of which 70 (77%) assessed daily oral PrEP [4, 47, 49, 53, 54, 57, 58, 61–66, 68, 69, 74–79, 82, 85, 87, 89, 92, 97–101, 103, 104, 107–109, 111–115, 117, 118, 124, 126–133, 135, 136, 139–146, 149, 151–156, 158], 12 (13%) assessed on-demand oral PrEP [72, 76, 78, 94, 95, 101, 113, 115, 130, 145, 151, 158], and 18 (20%) assessed oral PrEP without specifying daily oral or on-demand [48, 51, 52, 56, 80, 81, 86, 88, 91, 96, 102, 105, 116, 121, 123, 125, 137, 157]. Twelve (11%) of the 114 included studies evaluated long-acting injectable PrEP [74, 84, 100, 110, 125, 133, 135, 139, 145, 149, 155, 158]. Seven (6%) studies evaluated other PrEP modalities (e.g., vaginal ring, topical gel) [46, 50, 52, 59, 100, 106, 111]. Seventeen (15%) studies did not specify the PrEP modality (i.e., oral, injectable, vaginal ring, topical gel) evaluated [55, 60, 67, 70, 71, 73, 83, 90, 93, 119, 120, 122, 134, 138, 147, 148, 150].

### Type of comparators used over time in economic evaluations of PrEP

Of the 108 studies that included at least one HIV prevention intervention in the comparator, the most common comparator was a combination of ART and other HIV prevention interventions excluding PrEP (n=71; 66%; Figure 4) [51–55, 57–70, 73–75, 78–82, 84–86, 88, 91, 93, 94, 96, 98, 99, 103, 105–107, 109, 110, 112–120, 123–125, 127, 129–132, 135, 137, 141–146, 148, 152, 156]. Sixteen studies (15%), published between 2008 and 2024, only included ART in the comparator (i.e. no other concomitant HIV prevention interventions) [47–49, 54, 71, 76, 77, 83, 90, 92, 97, 104, 128, 140, 150, 154]. Three (3%) studies published from 2017 to 2019 did not include ART but included other concomitant HIV prevention interventions (e.g., condoms, HIV testing, HIV monitoring, and/or male circumcision) in their comparator [87, 100, 108]. One of the studies simulated an HIV epidemic between 2009 and 2019 [108] while the other two studies did not report the time period of the HIV epidemics simulated [87, 100]. Studies (n=9, 8%) that included PrEP along with other HIV prevention interventions in the comparator were all published in 2017 or later [89, 102, 122, 133, 134, 136, 147, 157, 158].

**Figure 4.**
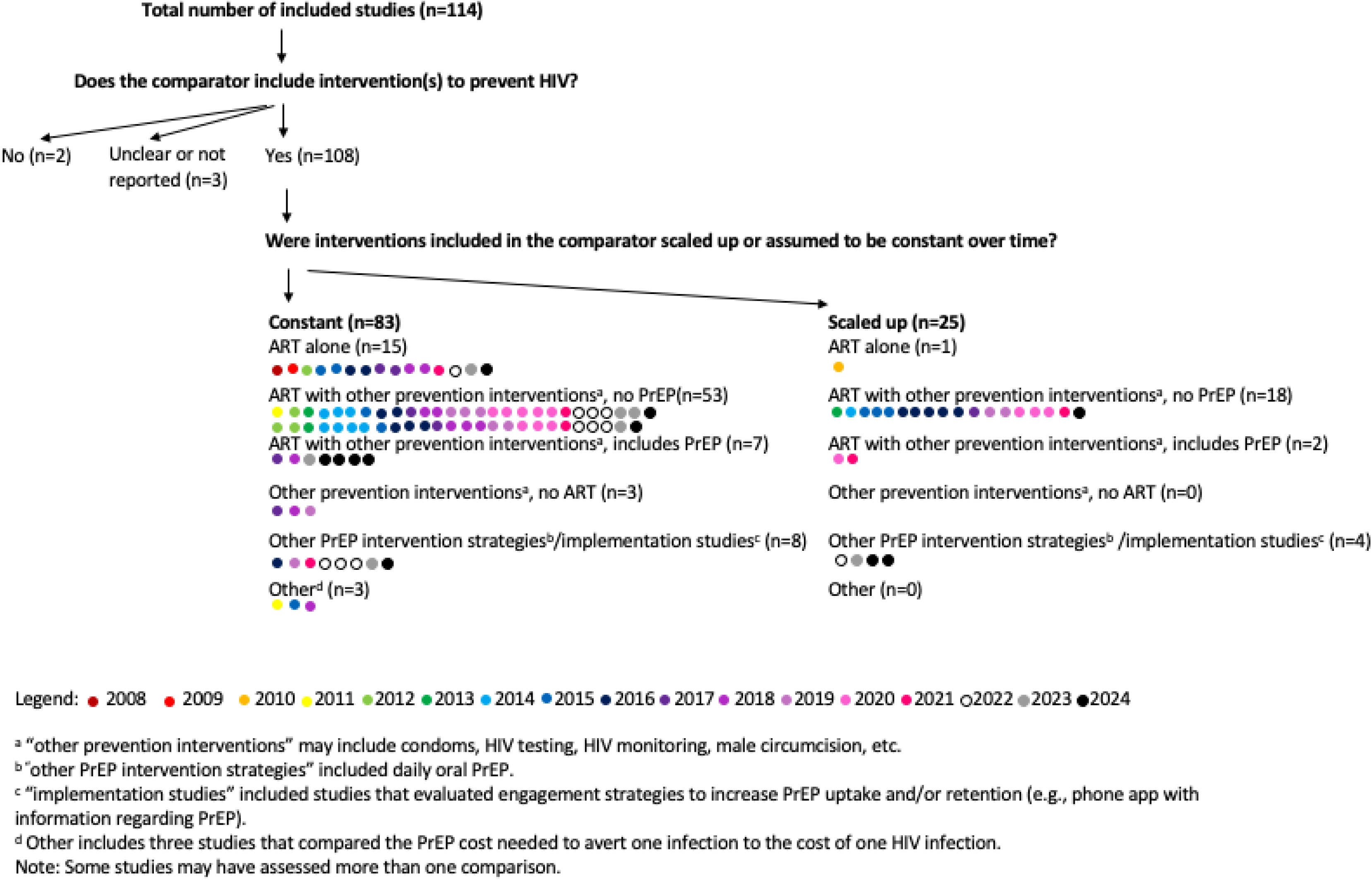
Types of comparators among included studies by applicability to real-world practice and year of study publication.

Fifty-three of the 71 studies (75%) that compared PrEP to ART and other HIV prevention interventions excluding PrEP assumed that the coverage of the comparators remained stable over time [51–54, 57–59, 61–66, 69, 70, 74, 78, 79, 82, 85, 88, 93, 94, 96, 98, 99, 103, 105, 107, 109, 112–119, 125, 127, 129, 130, 132, 135, 141–146, 148, 152, 156]. Of the 108 studies that included at least one HIV prevention intervention in the comparator, 22 (20%) considered the scale-up (i.e., increased coverage and increased uptake) of ART [49, 55, 60, 67, 68, 73, 75, 80, 81, 84, 86, 91, 106, 110, 120, 122–124, 131, 137, 147, 149], and 17 studies (16%) considered increased coverage and/or uptake of other prevention interventions over time [55, 60, 67, 68, 75, 80, 81, 84, 86, 91, 106, 122, 123, 137, 147, 151, 155].

### Economic evaluations comparing PrEP modalities and implementation strategies

Ten studies (9%) evaluated the cost-effectiveness of one PrEP modality versus another PrEP modality (Figure 4) including: 1) seven studies that compared long-acting injectable PrEP to daily or on-demand oral PrEP and other concomitant interventions (published between 2016 and 2024) [74, 111, 135, 139, 145, 149, 155]; 2) a study that compared dapivirine vaginal rings to daily oral PrEP with concomitant ART, condom use, and voluntary male circumcision (published in 2019) [111]; and 3) two studies that compared branded to generic daily oral PrEP without any concomitant interventions in the comparator (published in 2020) [126, 151].

Only three studies (3%), published between 2021 and 2024, examined the cost-effectiveness of different PrEP implementation strategies [4, 138, 153]. Three studies evaluated the cost-effectiveness of PrEP with and without PrEP engagement interventions (i.e., interventions to improve PrEP initiation, adherence, and persistence). PrEP initiation interventions assessed included: 1) a phone app with information about PrEP including individualized risk assessments and PrEP provider locations [4]; and 2) a phone app that assessed PrEP eligibility among gay, bisexual, and other men who have sex with men, and allowed users to order HIV/STI tests and safe sex products [153]. PrEP adherence interventions assessed included: 1) counseling [4, 153]; 2) text message support [138]; 3) PrEP patient navigation via trained healthcare professionals and navigators; and 4) educational training and PrEP screening questionnaires to help physicians integrate PrEP consultation and referral into visits [138]. Two studies evaluated the use of mobile phone apps that sought to facilitate telemedicine visits with PrEP providers to improve PrEP persistence [4, 153].

## Discussion

Our review identified 114 economic evaluations of PrEP, highlighting a rapidly growing number of studies over time. It highlights key areas for future work in assessing the cost-effectiveness of PrEP policy, programmes, and person-centered decisions. Almost three-quarters of published economic evaluations of PrEP examined HIV epidemics in 2015 or later, following WHO endorsement of PrEP and the increasing number of countries who have authorized the use of PrEP [7]. We identified a paucity of studies in the following areas: quantifying value for money from a broader societal perspective; capturing diverse epidemic context (geographic regions and populations); and evaluating different PrEP modalities and implementation strategies. Finally, a key gap across the current landscape of PrEP economic evaluations was the comparator: only 25 of the 114 studies included at least one HIV prevention intervention in the comparator and accounted for the concurrent scale-up of other HIV prevention interventions, reflecting current HIV prevention programmes.

Emerging PrEP modalities, particularly long-acting regimens, introduce costs (healthcare, transportation, accommodation, caregiving, productivity loss) and benefits (PrEP uptake and adherence, prevention of HIV transmission) that differ from existing or no PrEP scenarios [139]. These differences underscore the need to expand the perspective of economic evaluations to include societal impacts, capturing both immediate and downstream health benefits and quality of life as well as broader societal gains such as improved productivity and the spillover effects of improved public health [161, 162]. Previous costing studies have shown that PrEP can enhance work productivity by reducing the risk of HIV transmission [161, 163, 164]. Economic evaluations that only consider health system perspectives may underestimate PrEP’s value for money [165], potentially leading to suboptimal resource allocation decisions. As future economic analyses of PrEP modalities and/or PrEP implementation are developed, adopting a societal perspective would align more directly with the current era of an expansive choice in PrEP modalities, thereby supporting decision-making surrounding “person-centered” HIV prevention [166].

Our findings suggest important areas for future economic evaluations to keep pace with the evolving HIV response. Chief among them is the examination of different implementation strategies and PrEP modalities – particularly long-acting PrEP [167, 168]. Programmatic and policy decisions have shifted from the “technology” (i.e., whether PrEP should be offered), particularly among populations experiencing disproportionate risk, to implementation in an era of choice, with a focus on long-acting PrEP. Implementation includes focusing on how to reach those who would benefit at an individual-level or those who may be part of sexual and/or injecting networks so that PrEP use can reduce population-level HIV transmission as well as how to support user choices across regimens (building upon previous binary comparisons of oral PrEP vs. long-acting PrEP) [139]. Despite the expansion and availability of different PrEP modalities in the last decade and their inclusion in national guidelines [8, 9], PrEP coverage [10, 169], uptake [169], and adherence [170] remain programmatic challenges in many regions. In 2020, PrEP coverage was estimated at 8% of the estimated global need, and 28% of the estimated need in low- and middle-income countries [169]; with inequities in PrEP coverage by race, age, geography, and socioeconomic status [171]. As empirical data on the comparative reach and effectiveness of implementation strategies emerge, economic evaluations of implementation strategies delivering on choice can support the selection and scale-up of effective and efficient programmes. These include, but are not limited to, estimating the maximum pricing of PrEP and the pricing of programme/service delivery, for PrEP programmes to be considered good value for money by a country’s health system [172].

Alongside expansion of PrEP modalities, is the integration of HIV prevention services and programmes within a broader mandate of universal health care and service provision, and evolving epidemic response and epidemic dynamics surrounding populations at highest risk of new HIV infections [173]. Across many priority populations, HIV prevention programmes are part of community-based programmes [174]. In many countries, particularly in the global south, population-focused HIV prevention programmes may be integrated within the larger health sector, such that PrEP implementation strategies may change (along with cost and effectiveness of such strategies) [175]. A key gap identified in our review is the use of more relevant comparators – particularly the concomitant scale-up of combination HIV prevention including HIV treatment [38], which could tip the balance on cost-effectiveness analyses in either direction. While our review found that a range of populations have been considered in PrEP economic evaluations, most focussed on a range of underlying HIV risks among gay, bisexual, and other men who have sex with men. Thus, there remain subsets of the population at disproportionate HIV risk among whom future work could be conducted to support decision-making given heterogeneity in HIV risks and service delivery (programme implementation): including but not limited to individuals engaged in sex work (formal and informal); men who pay for sex (clients of sex workers); people who inject drug; adolescents and young women; transgender women; [176, 177]. Ensuring PrEP (across its modalities) is accessible to and reaches those who could benefit also means considering health equity as one of the population-level goals (in addition to reduction in population-level transmission). Such goals would benefit from integrating efforts at examining distributional cost-effectiveness and equity-informed economic evaluations [178]. Taken together, there remains an important opportunity for future economic evaluations to build on the prior literature, with a focus on likely programmatic shifts, current prevention gap, and comparators that mirror the current and potential trajectory (scale-up) of concomitant combination HIV prevention coverage over time.

## Limitations

Our study had several limitations. First, our review was restricted to peer-reviewed publications and did not include gray literature, such as reports from health technology assessment agencies, which often inform health system decisions. As such, our study may not fully represent the breadth of economic evaluations conducted, potentially overlooking some data used in policy decision-making. Second, the lack of detailed reporting in some studies hindered our ability to extract all variables of interest, resulting in gaps in our synthesis. Although reporting guidelines for full economic evaluations exist and their adoption is increasing [179, 180], many studies included in our review predate these guidelines. Consequently, these studies often omitted several key elements necessary for assessing methodological quality. This deficiency raises concerns about the reliability of their findings. Future work, as proposed, includes conducting a more detailed methodological review to assess the validity and reliability of included studies [42].

## Conclusions

Economic evaluations of HIV PrEP have grown over time. There remain important areas for future economic evaluations to prioritize, including consideration of societal costs and benefits, the broader impacts of PrEP, choice across a growing expanse of PrEP modalities, comparison across PrEP implementation strategies, and the most relevant comparators as the HIV epidemic dynamics evolve. The global HIV response is at a critical juncture, with integration into the universal health care mandate, overall reduction in funding and resources, sustainability and acceleration to end HIV as a public health threat, and a re-centering of health equity given persistent disproportionate risks among populations experiencing social, economic and structural marginalization [173, 181]. Programmatic and policy decisions that use estimates of value for money will need economic evaluations that mirror the current state of HIV prevention implementation and HIV epidemic realities and those anticipated into the near future.

## Data Availability

All data produced in the present work are contained in the manuscript.

## Acknowledgments

We thank helpful discussions with Nasheed Moqueet (Unity Health Toronto), Anna Simkin (Unity Health Toronto), and Olivia D’Silva (Ottawa Hospital Research Institute). SM’s research program is supported by a Tier 2 Canada Research Chair in Mathematical Modeling and Program Science (grant number 950-232643). DHST’s research program is supported by a Tier 2 Canada Research Chair in HIV Prevention and STI Research. Stefan Baral’s effort is supported by the Center for HIV and Mental Health Stigma Elimination Strategies (CHIMES), funded by the National Institutes of Mental Health (P30MH136919). SB’s effort is supported by the Center for HIV and Mental Health Stigma Elimination Strategies (CHIMES), funded by the National Institutes of Mental Health (P30MH136919).

