## Appendices A and B for "Evolving landscape of economic evaluations of HIV pre-exposure prophylaxis and pre-exposure prophylaxis implementation strategies: A systematic review"

**Appendix A. PRISMA Checklist**

| **Section and Topic** | **Item #** | **Checklist item** | **Location where item is reported** |
| --- | --- | --- | --- |
| **TITLE** | | |  |
| Title | 1 | Identify the report as a systematic review. | Page 1 |
| **ABSTRACT** | | |  |
| Abstract | 2 | See the PRISMA 2020 for Abstracts checklist. | Pages 3-4 |
| **INTRODUCTION** | | |  |
| Rationale | 3 | Describe the rationale for the review in the context of existing knowledge. | Pages 5-7 |
| Objectives | 4 | Provide an explicit statement of the objective(s) or question(s) the review addresses. | Page 7 |
| **METHODS** | | |  |
| Eligibility criteria | 5 | Specify the inclusion and exclusion criteria for the review and how studies were grouped for the syntheses. | Pages 8-11 |
| Information sources | 6 | Specify all databases, registers, websites, organisations, reference lists and other sources searched or consulted to identify studies. Specify the date when each source was last searched or consulted. | Page 8 |
| Search strategy | 7 | Present the full search strategies for all databases, registers and websites, including any filters and limits used. | Appendix B |
| Selection process | 8 | Specify the methods used to decide whether a study met the inclusion criteria of the review, including how many reviewers screened each record and each report retrieved, whether they worked independently, and if applicable, details of automation tools used in the process. | Pages 8-9 |
| Data collection process | 9 | Specify the methods used to collect data from reports, including how many reviewers collected data from each report, whether they worked independently, any processes for obtaining or confirming data from study investigators, and if applicable, details of automation tools used in the process. | Pages 9-10 |
| Data items | 10a | List and define all outcomes for which data were sought. Specify whether all results that were compatible with each outcome domain in each study were sought (e.g. for all measures, time points, analyses), and if not, the methods used to decide which results to collect. | Pages 9-10 |
|  | 10b | List and define all other variables for which data were sought (e.g. participant and intervention characteristics, funding sources). Describe any assumptions made about any missing or unclear information. | Pages 9-10 |
| Study risk of bias assessment | 11 | Specify the methods used to assess risk of bias in the included studies, including details of the tool(s) used, how many reviewers assessed each study and whether they worked independently, and if applicable, details of automation tools used in the process. | NA |
| Effect measures | 12 | Specify for each outcome the effect measure(s) (e.g. risk ratio, mean difference) used in the synthesis or presentation of results. | NA |
| Synthesis methods | 13a | Describe the processes used to decide which studies were eligible for each synthesis (e.g. tabulating the study intervention characteristics and comparing against the planned groups for each synthesis (item #5)). | NA |
|  | 13b | Describe any methods required to prepare the data for presentation or synthesis, such as handling of missing summary statistics, or data conversions. | NA |
|  | 13c | Describe any methods used to tabulate or visually display results of individual studies and syntheses. | Pages 10-11 |
|  | 13d | Describe any methods used to synthesize results and provide a rationale for the choice(s). If meta-analysis was performed, describe the model(s), method(s) to identify the presence and extent of statistical heterogeneity, and software package(s) used. | Pages 10-11 |
|  | 13e | Describe any methods used to explore possible causes of heterogeneity among study results (e.g. subgroup analysis, meta-regression). | NA |
|  | 13f | Describe any sensitivity analyses conducted to assess robustness of the synthesized results. | NA |
| Reporting bias assessment | 14 | Describe any methods used to assess risk of bias due to missing results in a synthesis (arising from reporting biases). | NA |
| Certainty assessment | 15 | Describe any methods used to assess certainty (or confidence) in the body of evidence for an outcome. | NA |
| **RESULTS** | | |  |
| Study selection | 16a | Describe the results of the search and selection process, from the number of records identified in the search to the number of studies included in the review, ideally using a flow diagram. | Page 11 |
|  | 16b | Cite studies that might appear to meet the inclusion criteria, but which were excluded, and explain why they were excluded. | NA |
| Study characteristics | 17 | Cite each included study and present its characteristics. | Pages 11-17 |
| Risk of bias in studies | 18 | Present assessments of risk of bias for each included study. | NA |
| Results of individual studies | 19 | For all outcomes, present, for each study: (a) summary statistics for each group (where appropriate) and (b) an effect estimate and its precision (e.g. confidence/credible interval), ideally using structured tables or plots. | NA |
| Results of syntheses | 20a | For each synthesis, briefly summarise the characteristics and risk of bias among contributing studies. | NA |
|  | 20b | Present results of all statistical syntheses conducted. If meta-analysis was done, present for each the summary estimate and its precision (e.g. confidence/credible interval) and measures of statistical heterogeneity. If comparing groups, describe the direction of the effect. | NA |
|  | 20c | Present results of all investigations of possible causes of heterogeneity among study results. | NA |
|  | 20d | Present results of all sensitivity analyses conducted to assess the robustness of the synthesized results. | NA |
| Reporting biases | 21 | Present assessments of risk of bias due to missing results (arising from reporting biases) for each synthesis assessed. | NA |
| Certainty of evidence | 22 | Present assessments of certainty (or confidence) in the body of evidence for each outcome assessed. | NA |
| **DISCUSSION** | | |  |
| Discussion | 23a | Provide a general interpretation of the results in the context of other evidence. | Pages 17-21 |
|  | 23b | Discuss any limitations of the evidence included in the review. | Pages 17-21 |
|  | 23c | Discuss any limitations of the review processes used. | Pages 20-21 |
|  | 23d | Discuss implications of the results for practice, policy, and future research. | Pages 17-21 |
| **OTHER INFORMATION** | | |  |
| Registration and protocol | 24a | Provide registration information for the review, including register name and registration number, or state that the review was not registered. | Page 7 |
|  | 24b | Indicate where the review protocol can be accessed, or state that a protocol was not prepared. | Pages 7-8 |
|  | 24c | Describe and explain any amendments to information provided at registration or in the protocol. | Pages 7-8 |
| Support | 25 | Describe sources of financial or non-financial support for the review, and the role of the funders or sponsors in the review. | Page 27 |
| Competing interests | 26 | Declare any competing interests of review authors. | Page 27 |
| Availability of data, code and other materials | 27 | Report which of the following are publicly available and where they can be found: template data collection forms; data extracted from included studies; data used for all analyses; analytic code; any other materials used in the review. | Appendices |

*From:* Page MJ, McKenzie JE, Bossuyt PM, Boutron I, Hoffmann TC, Mulrow CD, et al. The PRISMA 2020 statement: an updated guideline for reporting systematic reviews. BMJ 2021;372:n71. doi: 10.1136/bmj.n71. This work is licensed under CC BY 4.0.

**Appendix B. Search Strategy**

HIV PrEP – Econ

Update

2024 Dec 21

*Updated from 2021 Dec 23*

*Updated from 2019 Aug 30*

Ovid Multifile

Database: Embase Classic+Embase <1947 to 2024 December 20>, Ovid MEDLINE(R) ALL <1946 to December 20, 2024>

Search Strategy:

--------------------------------------------------------------------------------

1	HIV Seronegativity/ (53624)

2	((AIDs or (acquired immune deficiency* adj2 virus) or (acquired immunodeficiency* adj2 virus*) or human immunodeficiency virus* or human immuno-deficiency virus*) adj2 (negativ* or seronegativ* or sero-negativ*)).tw,kf. (2824)

3	((HIV or HIV-1 or HIV1 or HIV-I or HIVI or HIV-2 or HIV2 or HIV-2 or HIV-II or HIVII or HTLV* or LAV-2 or LAV-II) adj2 (negativ* or seronegativ* or sero-negativ*)).tw,kf. (43190)

4	((uninfected or "not infected") adj2 (HIV or HIV-1 or HIV1 or HIV-I or HIVI or HIV-2 or HIV2 or HIV-2 or HIV-II or HIVII or HTLV* or LAV-2 or LAV-II)).tw,kf. (11891)

5	((uninfected or "not infected") adj2 (AIDs or (acquired immune deficiency* adj2 virus) or (acquired immunodeficiency* adj2 virus*) or human immunodeficiency virus* or human immuno-deficiency virus*)).tw,kf. (455)

6	(without adj (HIV or HIV-1 or HIV1 or HIV-I or HIVI or HIV-2 or HIV2 or HIV-2 or HIV-II or HIVII or HTLV* or LAV-2 or LAV-II)).tw,kf. (6309)

7	(without adj (AIDs or (acquired immune deficiency* adj2 virus) or (acquired immunodeficiency* adj2 virus*) or human immunodeficiency virus* or human immuno-deficiency virus*)).tw,kf. (2523)

8	((non or "not") adj (HIV or HIV-1 or HIV1 or HIV-I or HIVI or HIV-2 or HIV2 or HIV-2 or HIV-II or HIVII or HTLV* or LAV-2 or LAV-II) adj positiv*).tw,kf. (279)

9	(non-positiv* adj (HIV or HIV-1 or HIV1 or HIV-I or HIVI or HIV-2 or HIV2 or HIV-2 or HIV-II or HIVII or HTLV* or LAV-2 or LAV-II)).tw,kf. (0)

10	((non or "not") adj (AIDs or (acquired immune deficiency* adj2 virus) or (acquired immunodeficiency* adj2 virus*) or human immunodeficiency virus* or human immuno-deficiency virus*) adj positiv*).tw,kf. (2)

11	(non-positiv* adj (AIDs or (acquired immune deficiency* adj2 virus) or (acquired immunodeficiency* adj2 virus*) or human immunodeficiency virus* or human immuno-deficiency virus*)).tw,kf. (0)

12	or/1-11 [HIV SERONEGATIVITY] (112027)

13	exp HIV Infections/pc [Prevention & Control] (101417)

14	(prevent* adj3 (AIDs or (acquired immune deficiency* adj2 virus) or (acquired immunodeficiency* adj2 virus*) or human immunodeficiency virus* or human immuno-deficiency virus*)).tw,kf. (12346)

15	(prevent* adj3 (HIV or HIV-1 or HIV1 or HIV-I or HIVI or HIV-2 or HIV2 or HIV-2 or HIV-II or HIVII or HTLV* or LAV-2 or LAV-II)).tw,kf. (57581)

16	(prophyla* adj3 (AIDs or (acquired immune deficiency* adj2 virus) or (acquired immunodeficiency* adj2 virus*) or human immunodeficiency virus* or human immuno-deficiency virus*)).tw,kf. (1010)

17	(prophyla* adj3 (HIV or HIV-1 or HIV1 or HIV-I or HIVI or HIV-2 or HIV2 or HIV-2 or HIV-II or HIVII or HTLV* or LAV-2 or LAV-II)).tw,kf. (7752)

18	or/13-17 [HIV PROPHYLAXIS] (138016)

19	Anti-Retroviral Agents/ (52087)

20	Anti-HIV Agents/ (61705)

21	(antiretrovir* or anti-retrovir* or antiHIV* or anti-HIV* or antiAIDS or anti-AIDS).tw,kf. (192614)

22	Adenine/aa (9728)

23	Tenofovir/ or tenofovir*.tw,kf. (33669)

24	107021-12-5.rn. (0)

25	Rilpivirine/ or (rilpivirine or edurant or TMC-278 or TMC278 or UNII-FI96A8X663).tw,kf. (3786)

26	rilpivirine.rn. (2847)

27	(dapivirine or TMC-120 or TMC120 or UNII-TCN4MG2VXS).tw,kf. (789)

28	dapivirine.rn. (516)

29	"Emtricitabine, Tenofovir Disoproxil Fumarate Drug Combination"/ or Truvada.tw,kf. (4880)

30	("TDF-FTC" or "FTC-TDF" or "TDF/FTC" or "FTC/TDF").tw,kf. (2534)

31	731772-45-5.rn. (562)

32	or/19-31 [ANTI-HIV/RETROVIRAL AGENTS] (263215)

33	Primary Prevention/ or Pre-Exposure Prophylaxis/ (73270)

34	32 and 33 (5407)

35	12 and (18 or 34) (9718)

36	18 and 32 (29180)

37	exp HIV Infections/ (700723)

38	exp HIV/ (311244)

39	HIV Seroprevalence/ (13837)

40	(AIDs or (acquired immune deficiency* adj2 virus) or (acquired immunodeficiency* adj2 virus*) or human immunodeficiency virus* or human immuno-deficiency virus*).tw,kf. (519030)

41	(HIV or HIV-1 or HIV1 or HIV-I or HIVI or HIV-2 or HIV2 or HIV-2 or HIV-II or HIVII or HTLV* or LAV-2 or LAV-II).tw,kf. (790856)

42	or/37-41 [HIV] (1136370)

43	Pre-Exposure Prophylaxis/ (10088)

44	((pre-expos* or preexpos*) adj3 prophyla*).tw,kf. (11574)

45	((pre-expos* or preexpos*) adj3 protect*).tw,kf. (219)

46	PrEP.tw,kf. (18959)

47	or/43-46 [PrEP] (24260)

48	(1 or 42) and 47 (13540)

49	35 or 36 or 48 (42186)

50	Economics/ (273229)

51	exp "Costs and Cost Analysis"/ (629811)

52	Economics, Nursing/ (40883)

53	Economics, Medical/ (46942)

54	Economics, Pharmaceutical/ (11763)

55	exp Economics, Hospital/ (981558)

56	Economics, Dental/ (40661)

57	exp "Fees and Charges"/ (74694)

58	exp Budgets/ (45498)

59	budget*.ti,ab. (75492)

60	(economic* or cost or costs or costly or costing or price or prices or pricing or pharmacoeconomic* or pharmaco-economic* or expenditure or expenditures or expense or expenses or financial or finance or finances or financed).ti. (485325)

61	(economic* or cost or costs or costly or costing or price or prices or pricing or pharmacoeconomic* or pharmaco-economic* or expenditure or expenditures or expense or expenses or financial or finance or finances or financed).ab. /freq=2 (800538)

62	(cost* adj2 (effective* or utilit* or benefit* or minimi* or analy* or outcome or outcomes)).ab. (431306)

63	(value adj2 (money or monetary)).ti,ab. (6369)

64	exp Models, Economic/ (18779)

65	Quality-Adjusted Life Years/ (44536)

66	(life qualities or life quality or quality adjusted or adjusted life or qol or qoly or qolys or hrqol or qaly or qalys or qale or qales).tw. (247929)

67	exp Models, Economic/ (18779)

68	economic model*.ti,ab. (9339)

69	agent-based.ti,ab. (9236)

70	individual-based.ti,ab. (7996)

71	transmission dynamic?.ti,ab. (9376)

72	deterministic.ti,ab. (36123)

73	compartmental.ti,ab. (26281)

74	(compartment* adj2 model*).ti,ab. (46374)

75	(discrete event? adj2 (model* or simulat*)).ti,ab. (2430)

76	ordinary differential equation?.ti,ab. (7159)

77	ode model*.ti,ab. (819)

78	Stochastic Processes/ (33202)

79	stochastic.ti,ab. (87439)

80	Markov Chains/ (23244)

81	markov.ti,ab. (56538)

82	Monte Carlo Method/ (75568)

83	monte carlo.ti,ab. (107339)

84	exp Decision Theory/ (14567)

85	(decision* adj2 (tree* or analy* or model*)).ti,ab. (69056)

86	or/50-85 (2751669)

87	49 and 86 (5237)

88	"Quality of Life"/ (764098)

89	Quality-Adjusted Life Years/ (44536)

90	(life adj1 (quality or qualities)).ti,ab. (25724)

91	(adjusted adj1 (quality or life)).ti,ab. (49360)

92	(qol or qoly or qolys or hrqol or qaly or qalys or qale or qales).ti,ab. (206818)

93	or/88-92 (856593)

94	49 and 93 (762)

95	87 or 94 (5477)

96	exp Animals/ not (exp Animals/ and Humans/) (17944991)

97	95 not 96 [ANIMAL-ONLY REMOVED] (4178)

98	(20190829* or 2019083* or 201909* or 201910* or 201911* or 201912* or 2020* or 2021*).dt. (3439829)

99	97 and 98 [MEDLINE UPDATE PERIOD] (243)

100	99 use medall [MEDLINE RECORDS] (243)

101	((AIDs or (acquired immune deficiency* adj2 virus) or (acquired immunodeficiency* adj2 virus*) or human immunodeficiency virus* or human immuno-deficiency virus*) adj2 (negativ* or seronegativ* or sero-negativ*)).tw,kw. (2832)

102	((HIV or HIV-1 or HIV1 or HIV-I or HIVI or HIV-2 or HIV2 or HIV-2 or HIV-II or HIVII or HTLV* or LAV-2 or LAV-II) adj2 (negativ* or seronegativ* or sero-negativ*)).tw,kw. (43144)

103	((uninfected or "not infected") adj2 (HIV or HIV-1 or HIV1 or HIV-I or HIVI or HIV-2 or HIV2 or HIV-2 or HIV-II or HIVII or HTLV* or LAV-2 or LAV-II)).tw,kw. (11851)

104	((uninfected or "not infected") adj2 (AIDs or (acquired immune deficiency* adj2 virus) or (acquired immunodeficiency* adj2 virus*) or human immunodeficiency virus* or human immuno-deficiency virus*)).tw,kw. (454)

105	(without adj (HIV or HIV-1 or HIV1 or HIV-I or HIVI or HIV-2 or HIV2 or HIV-2 or HIV-II or HIVII or HTLV* or LAV-2 or LAV-II)).tw,kw. (6306)

106	(without adj (AIDs or (acquired immune deficiency* adj2 virus) or (acquired immunodeficiency* adj2 virus*) or human immunodeficiency virus* or human immuno-deficiency virus*)).tw,kw. (2523)

107	((non or "not") adj (HIV or HIV-1 or HIV1 or HIV-I or HIVI or HIV-2 or HIV2 or HIV-2 or HIV-II or HIVII or HTLV* or LAV-2 or LAV-II) adj positiv*).tw,kw. (279)

108	(non-positiv* adj (HIV or HIV-1 or HIV1 or HIV-I or HIVI or HIV-2 or HIV2 or HIV-2 or HIV-II or HIVII or HTLV* or LAV-2 or LAV-II)).tw,kw. (0)

109	((non or "not") adj (AIDs or (acquired immune deficiency* adj2 virus) or (acquired immunodeficiency* adj2 virus*) or human immunodeficiency virus* or human immuno-deficiency virus*) adj positiv*).tw,kw. (2)

110	(non-positiv* adj (AIDs or (acquired immune deficiency* adj2 virus) or (acquired immunodeficiency* adj2 virus*) or human immunodeficiency virus* or human immuno-deficiency virus*)).tw,kw. (0)

111	or/101-110 [HIV SEROPOSITIVITY] (62335)

112	exp Human immunodeficiency virus infection/pc [Prevention] (48311)

113	(prevent* adj3 (AIDs or (acquired immune deficiency* adj2 virus) or (acquired immunodeficiency* adj2 virus*) or human immunodeficiency virus* or human immuno-deficiency virus*)).tw,kw. (12647)

114	(prevent* adj3 (HIV or HIV-1 or HIV1 or HIV-I or HIVI or HIV-2 or HIV2 or HIV-2 or HIV-II or HIVII or HTLV* or LAV-2 or LAV-II)).tw,kw. (55887)

115	(prophyla* adj3 (AIDs or (acquired immune deficiency* adj2 virus) or (acquired immunodeficiency* adj2 virus*) or human immunodeficiency virus* or human immuno-deficiency virus*)).tw,kw. (1119)

116	(prophyla* adj3 (HIV or HIV-1 or HIV1 or HIV-I or HIVI or HIV-2 or HIV2 or HIV-2 or HIV-II or HIVII or HTLV* or LAV-2 or LAV-II)).tw,kw. (7803)

117	or/112-116 [HIV PROPHYLAXIS] (101337)

118	antiretrovirus agent/ (43138)

119	anti human immunodeficiency virus agent/ (21597)

120	(antiretrovir* or anti-retrovir* or antiHIV* or anti-HIV* or antiAIDS or anti-AIDS).tw,kw. (192144)

121	tenofovir/ (23927)

122	tenofovir.rn. (21412)

123	rilpivirine/ (3098)

124	rilpivirine.rn. (2847)

125	dapivirine/ (736)

126	dapivirine.rn. (516)

127	emtricitabine plus tenofovir disoproxil/ (4330)

128	Truvada.tw,kw. (1943)

129	("TDF-FTC" or "FTC-TDF" or "TDF/FTC" or "FTC/TDF").tw,kw. (2533)

130	731772-45-5.rn. (562)

131	or/118-130 [ANTI-HIV/RETROVIRAL AGENTS] (235151)

132	prophylaxis/ or pre-exposure prophylaxis/ (142660)

133	131 and 132 (6162)

134	111 and (117 or 133) (7133)

135	117 and 131 (21703)

136	exp Human immunodeficiency virus infection/ (400677)

137	exp Human immunodeficiency virus/ (311244)

138	human immunodeficiency virus prevalence/ (10636)

139	(AIDs or (acquired immune deficiency* adj2 virus) or (acquired immunodeficiency* adj2 virus*) or human immunodeficiency virus* or human immuno-deficiency virus*).tw,kw. (507789)

140	(HIV or HIV-1 or HIV1 or HIV-I or HIVI or HIV-2 or HIV2 or HIV-2 or HIV-II or HIVII or HTLV* or LAV-2 or LAV-II).tw,kw. (786131)

141	or/136-140 [HIV] (1098044)

142	pre-exposure prophylaxis/ (10088)

143	((pre-expos* or preexpos*) adj3 prophyla*).tw,kw. (11146)

144	((pre-expos* or preexpos*) adj3 protect*).tw,kw. (219)

145	PrEP.tw,kw. (18710)

146	or/142-145 [PrEP] (23911)

147	141 and 146 [HIV PrEP] (13027)

148	134 or 135 or 147 [HIV PrEP] (34027)

149	economics/ (273229)

150	exp cost/ (629811)

151	exp health economics/ (2565506)

152	exp fee/ (74694)

153	budget/ (43097)

154	budget*.ti,ab. (75492)

155	(economic* or cost or costs or costly or costing or price or prices or pricing or pharmacoeconomic* or pharmaco-economic* or expenditure or expenditures or expense or expenses or financial or finance or finances or financed).ti. (485325)

156	(economic* or cost or costs or costly or costing or price or prices or pricing or pharmacoeconomic* or pharmaco-economic* or expenditure or expenditures or expense or expenses or financial or finance or finances or financed).ab. /freq=2 (800538)

157	(cost* adj2 (effective* or utilit* or benefit* or minimi* or analy* or outcome or outcomes)).ab. (431306)

158	(value adj2 (money or monetary)).ti,ab. (6369)

159	statistical model/ (265373)

160	economic model*.ti,ab. (9339)

161	probability/ (185382)

162	mathematical model/ (286910)

163	agent-based.ti,ab. (9236)

164	individual-based.ti,ab. (7996)

165	transmission dynamic?.ti,ab. (9376)

166	deterministic.ti,ab. (36123)

167	compartment model/ (17247)

168	compartmental.ti,ab. (26281)

169	(compartment* adj2 model*).ti,ab. (46374)

170	(discrete event? adj2 (model* or simulat*)).ti,ab. (2430)

171	ordinary differential equation?.ti,ab. (7159)

172	ode model*.ti,ab. (819)

173	stochastic model/ (18097)

174	stochastic.ti,ab. (87439)

175	markov.ti,ab. (56538)

176	monte carlo method/ (75568)

177	monte carlo.ti,ab. (107339)

178	decision theory/ (2789)

179	(decision* adj2 (tree* or analy* or model*)).ti,ab. (69056)

180	or/149-179 (4370336)

181	148 and 180 (5688)

182	exp "quality of life"/ (789408)

183	(life adj1 (quality or qualities)).ti,ab. (25724)

184	(adjusted adj1 (quality or life)).ti,ab. (49360)

185	(qol or qoly or qolys or hrqol or qaly or qalys or qale or qales).ti,ab. (206818)

186	or/182-185 (853612)

187	148 and 186 (668)

188	181 or 187 (5937)

189	exp animal experimentation/ or exp models animal/ or exp animal experiment/ or nonhuman/ or exp vertebrate/ (55676405)

190	exp human/ or exp human experimentation/ or exp human experiment/ (44414827)

191	189 not 190 (11263499)

192	188 not 191 [ANIMAL-ONLY REMOVED] (5897)

193	conference abstract.pt. (4280716)

194	192 not 193 [CONFERENCE ABSTRACTS REMOVED] (5363)

195	(20190829* or 2019083* or 201909* or 201910* or 201911* or 201912* or 2020* or 2021*).dc. (4662134)

196	194 and 195 [EMBASE UPDATE PERIOD] (543)

197	196 use emczd [EMBASE RECORDS] (543)

198	100 or 197 [BOTH DATABASES - UPDATE PERIOD] (786)

199	remove duplicates from 198 (592) [TOTAL UNIQUE RECORDS – UPDATE PERIOD]

200	199 use medall [MEDLINE UNIQUE RECORDS] (243)

201	199 use emczd [EMBASE UNIQUE RECORDS] (349)

***************************
